# Albumin–Neutrophil Composite Grading (ANPG) for Predicting Overall Survival in Colorectal Cancer: A Retrospective Cohort Study

**DOI:** 10.64898/2026.01.06.26343565

**Authors:** Xuelian Shi, Guo Tian, Chaoxi Zhou, Jiena Zhou, Haiyan Fu, Chunfu Wan, Xiaoli Xu

## Abstract

**Background:** Inflammatory responses and nutritional status are critical determinants of colorectal cancer (CRC) progression and clinical outcomes. This study aimed to evaluate the prognostic value of a novel albumin-neutrophil composite grading (ANPG) system and to develop a nomogram for predicting overall survival (OS) in CRC patients following curative resection.

**Methods:** A retrospective analysis was conducted on 660 consecutive patients with primary CRC who underwent R0 resection between December 2017 and December 2018. The ANPG was constructed based on preoperative serum albumin levels and neutrophil counts, with optimal cutoff values determined by receiver operating characteristic (ROC) curve analysis. Prognostic factors were identified using univariate and multivariate Cox proportional hazards regression models. A predictive nomogram was developed and internally validated via bootstrap resampling (800 iterations) and time-dependent ROC analysis. Decision curve analysis (DCA) was performed to assess clinical utility.

**Results:** The median follow-up duration was 2442 days (interquartile range: 2117–2537 days), during which 108 cancer-specific deaths occurred. The ANPG demonstrated superior discriminative ability (area under the curve [AUC] = 0.637, 95% confidence interval [CI]: 0.588–0.687, P < 0.001) compared to established inflammatory and nutritional markers, including the neutrophil-to-lymphocyte ratio (NLR), platelet-to-lymphocyte ratio (PLR), systemic immune-inflammation index (SII), fibrinogen-to-albumin ratio (FAR), and fibrinogen-neutrophil-lymphocyte ratio (F-NLR). Kaplan-Meier survival analysis revealed significant differences in OS across ANPG grades (Log-rank χ² = 24.423, P < 0.001), with 5-year OS rates of 93.7%, 83.2%, and 74.4% for grades 0, 1, and 2, respectively. Multivariate Cox regression analysis identified ANPG (grade 1 vs. 0: hazard ratio [HR] = 2.190, P = 0.020; grade 2 vs. 0: HR = 3.256, P < 0.001), age (HR = 1.032, P < 0.001), carbohydrate antigen 19-9 (CA19-9) (HR = 1.002, P = 0.003), histological type (HR = 1.954, P = 0.005), and TNM stage as independent prognostic factors. The nomogram incorporating these variables—retaining carcinoembryonic antigen due to its clinical relevance and contribution to model performance—achieved a concordance index (C-index) of 0.806 (95% CI: 0.788–0.824) with excellent calibration, significantly outperforming TNM staging alone in predictive accuracy. Decision curve analysis (DCA) showed greater net benefit across a wide range of threshold probabilities for 5-year OS compared to TNM staging, confirming its enhanced clinical applicability.

**Conclusions:** The ANPG is a robust and independent prognostic indicator in CRC patients undergoing curative resection. The developed nomogram provides a clinically valuable tool for individualized survival prediction and risk stratification.

## Introduction

Colorectal cancer (CRC) represents a major global health burden, ranking as the third most frequently diagnosed malignancy and the second leading cause of cancer-related mortality worldwide^[1]^. Despite advances in surgical techniques and adjuvant therapies, disease recurrence or metastasis develops in 20–30% of patients after curative-intent resection, highlighting the necessity for accurate prognostic assessment to guide treatment decisions and postoperative surveillance^[2, 3]^.

The tumor-node-metastasis (TNM) classification system remains the cornerstone of colorectal cancer (CRC) prognostic stratification. However, this anatomical staging approach has inherent limitations in predicting individual patient outcomes, as demonstrated by considerable heterogeneity in clinical progression among patients with identical TNM stages^[4–6]^. This highlights the need to incorporate additional prognostic markers to improve risk assessment accuracy.

Accumulating evidence suggests that cancer-related systemic inflammation and nutritional status play pivotal roles in disease progression and clinical outcomes^[7–9]^. The interplay between chronic inflammation and malnutrition contributes to a tumor-permissive microenvironment, promoting cellular proliferation, local invasion, and distant metastasis^[10, 11]^. Several inflammation– and nutrition-based prognostic scores have been evaluated in CRC, including the neutrophil-to-lymphocyte ratio (NLR), platelet-to-lymphocyte ratio (PLR), systemic immune-inflammation index (SII), fibrinogen-to-albumin ratio (FAR), and fibrinogen-neutrophil-lymphocyte ratio (F-NLR)^[12–18]^. However, these existing composite indices—largely derived from simplified ratios or limited parameter combinations (e.g., F-NLR)—fail to adequately capture the complex and integrated influence of inflammatory-nutritional interactions on CRC prognosis.

The albumin-neutrophil composite grading (ANPG) system represents a novel prognostic model that simultaneously assesses nutritional (albumin) and inflammatory (neutrophil) parameters. Serum albumin is a well-established marker of both nutritional status and systemic inflammation, with hypoalbuminemia consistently linked to poor outcomes across various malignancies^[19, 20]^. Neutrophils contribute to tumor-associated inflammation by releasing pro-tumorigenic mediators such as cytokines, chemokines, and reactive oxygen species, which promote tumor progression and immune evasion^[21, 22]^. By integrating these two clinically relevant parameters, the albumin-neutrophil composite grading (ANPG) system provides a more comprehensive assessment of host inflammatory-nutritional status than single biomarkers or existing composite scores. A key advantage of the albumin-neutrophil composite grading (ANPG) system is its reliance on routine preoperative laboratory measurements—serum albumin and neutrophil count—which are widely available, cost-effective, and easy to implement in clinical practice. This simplicity enhances its applicability compared to multi-parameter models (e.g., SII, F-NLR) that require complex calculations or multiple hematological variables, thus supporting broader clinical adoption.

Although previous studies have investigated the prognostic value of the albumin-to-neutrophil percentage ratio (ANPG) in non-small cell lung cancer^[23]^, its role in colorectal cancer remains insufficiently characterized. This study aims to: (1) systematically assess the prognostic utility of ANPG in CRC patients following radical resection; (2) quantitatively compare its predictive performance with conventional inflammatory and nutritional biomarkers; and (3) develop and validate a prognostic nomogram incorporating ANPG for individualized survival prediction.

## Materials and Methods

### Study Design and Patient Selection

This retrospective cohort study was conducted at the Fourth Hospital of Hebei Medical University, including consecutive patients who underwent curative-intent surgery for primary CRC between December 2017 and December 2018. This study was approved by the Ethics Committee of the Fourth Hospital of Hebei Medical University (approval No. 2025KS056) and conducted in accordance with the Declaration of Helsinki. Owing to its retrospective design, the requirement for informed consent was waived.

Eligibility criteria included: (1) histologically confirmed primary colorectal cancer (CRC), including adenocarcinoma or mucinous adenocarcinoma; (2) complete R0 resection with microscopically negative margins; (3) documented preoperative serum albumin and neutrophil counts measured within 7 days prior to surgery; (4) availability of comprehensive follow-up data; and (5) for stage IVA (M1a) patients, synchronous single-organ metastasis (hepatic or pulmonary) with R0 resection of both primary and metastatic lesions, absence of peritoneal or multi-organ dissemination, and completion of 6 months of standard adjuvant chemotherapy.

Exclusion criteria were: (1) stage IVB/IVC disease or non-R0 resection; (2) concomitant malignancies, significant cardiovascular comorbidities, or end-stage organ dysfunction; (3) major postoperative complications (e.g., anastomotic leak complicated by septic shock); (4) prior neoadjuvant therapy or use of medications known to affect albumin levels or hematological parameters (e.g., corticosteroids, nonsteroidal anti-inflammatory drugs); (5) death from non-oncologic causes or loss to follow-up.

### Data-access and privacy statement

To further ensure patient privacy, we clarify that all records were fully de-identified before analysis; the authors had no access to any information that could identify individual participants during or after data collection. The clinical dataset was extracted from the electronic medical record system on 15 January 2024 for research purposes. The requirement for informed consent was waived by the Ethics Committee of the Fourth Hospital of Hebei Medical University (approval No. 2025KS056).

### Data Collection and Variable Definition

Clinical and pathological variables were systematically extracted from electronic medical records, including demographic characteristics (age, sex), tumor-related features (TNM stage according to the 8th edition of the American Joint Committee on Cancer [AJCC] staging system, histological subtype, perineural invasion [PNI], lymphovascular invasion [LVI]), therapeutic interventions (postoperative adjuvant chemotherapy), and preoperative laboratory markers (serum albumin, neutrophil count, lymphocyte count, platelet count, fibrinogen, carcinoembryonic antigen [CEA], and carbohydrate antigen 19-9 [CA19-9]).

### The following inflammatory and nutritional indices were calculated

Neutrophil-to-lymphocyte ratio (NLR): defined as neutrophil count divided by lymphocyte count. Platelet-to-lymphocyte ratio (PLR): calculated as platelet count divided by lymphocyte count. Systemic immune-inflammation index (SII): computed as (platelet count × neutrophil count) / lymphocyte count. Fibrinogen-to-albumin ratio (FAR): determined as fibrinogen concentration divided by albumin concentration. Fibrinogen-neutrophil-lymphocyte ratio (F-NLR) score: patients were classified into three groups based on predefined cutoffs (fibrinogen ≥3.595 g/L and NLR ≥3.0157): score 2 (both elevated), score 1 (one elevated), or score 0 (neither elevated).

### Albumin-Neutrophil Composite Grading (ANPG) System

The ANPG system was developed using preoperative serum albumin and neutrophil count values. Optimal cutoffs were identified through receiver operating characteristic (ROC) curve analysis, with the maximum Youden index (sensitivity + specificity − 1) used to determine thresholds for predicting overall survival. The optimal cutoffs were 42.35 g/L for albumin and 3.755×10⁹/L for neutrophils.

### Patients were categorized into three ANPG grades

Grade 0 (well-nourished with mild inflammation): albumin ≥42.35 g/L and neutrophil count <3.755×10⁹/L. Grade 1 (single nutritional or inflammatory abnormality): either albumin ≥42.35 g/L and neutrophil count ≥3.755×10⁹/L (well-nourished with systemic inflammation), or albumin <42.35 g/L and neutrophil count <3.755×10⁹/L (malnourished without significant inflammation). Grade 2 (malnourished with severe inflammation): albumin <42.35 g/L and neutrophil count ≥3.755×10⁹/L.

### Follow-up and Study Endpoint

Postoperative follow-up was performed at 3–6-month intervals during the first 2 years and annually thereafter. Follow-up assessments included outpatient clinic visits, telephone interviews, and review of electronic medical records. The primary endpoint was overall survival (OS), defined as the time from date of surgery to date of death from any cause or last known follow-up. The study follow-up period ended on December 16, 2024.

### Statistical Analysis

Statistical analyses were performed using SPSS version 25.0 (IBM Corp., Armonk, NY, USA) and R version 4.2.1 (R Foundation for Statistical Computing, Vienna, Austria), with the following packages: survival (v3.3.1), rms (v6.3-0), pROC, and ggplot2. Continuous variables were summarized as median (interquartile range [IQR]) or mean ± standard deviation (SD), while categorical variables were presented as frequency (percentage). Receiver operating characteristic (ROC) curve analysis was used to assess the discriminative performance of biomarkers, with area under the curve (AUC) values and corresponding 95% confidence intervals (CIs) calculated. Comparison of AUCs between different markers was conducted using DeLong’s test.

Survival analysis was carried out using the Kaplan-Meier method, and group differences were evaluated by the log-rank test. Univariate and multivariate Cox proportional hazards regression models were applied to identify independent prognostic factors. Variables with a univariate P value < 0.10 were included in the multivariate model. Multicollinearity was assessed using the variance inflation factor (VIF), and a VIF < 4 was considered acceptable.

A nomogram was constructed based on the results of multivariate Cox regression using the rms package. Model discrimination was evaluated by the concordance index (C-index) and time-dependent ROC (tdROC) analysis. Calibration curves were generated to assess the agreement between predicted and observed survival probabilities, and internal validation was performed using bootstrap resampling (800 iterations, 100 samples per iteration). Clinical utility was further evaluated through decision curve analysis (DCA).

All statistical tests were two-sided, and a P value < 0.05 was considered statistically significant.

## Results

### Patient Characteristics

From December 2017 to December 2018, a total of 660 consecutive eligible patients were enrolled (Table 1). The median age was 61 years (IQR: 53–68), and 353 (53.5%) were male. TNM stage distribution was as follows: stage I (n = 102, 15.5%), stage II (n = 298, 45.2%), stage III (n = 249, 37.7%), and stage IVA (n = 11, 1.7%). The majority of tumors were conventional adenocarcinoma (n = 572, 86.7%), while mucinous adenocarcinoma accounted for 13.3% (n = 88). Postoperative adjuvant chemotherapy was administered to 407 patients (61.7%).

**Table 1:**
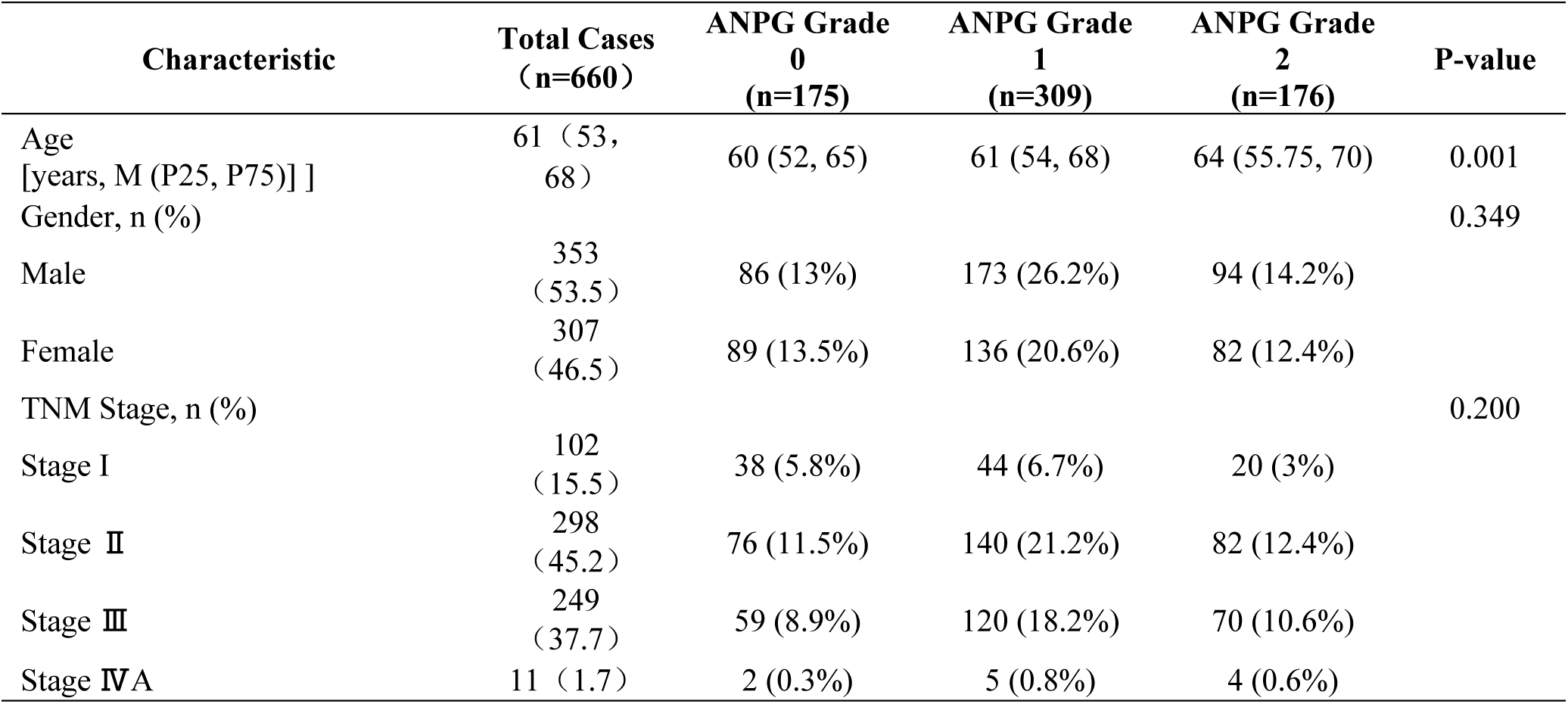
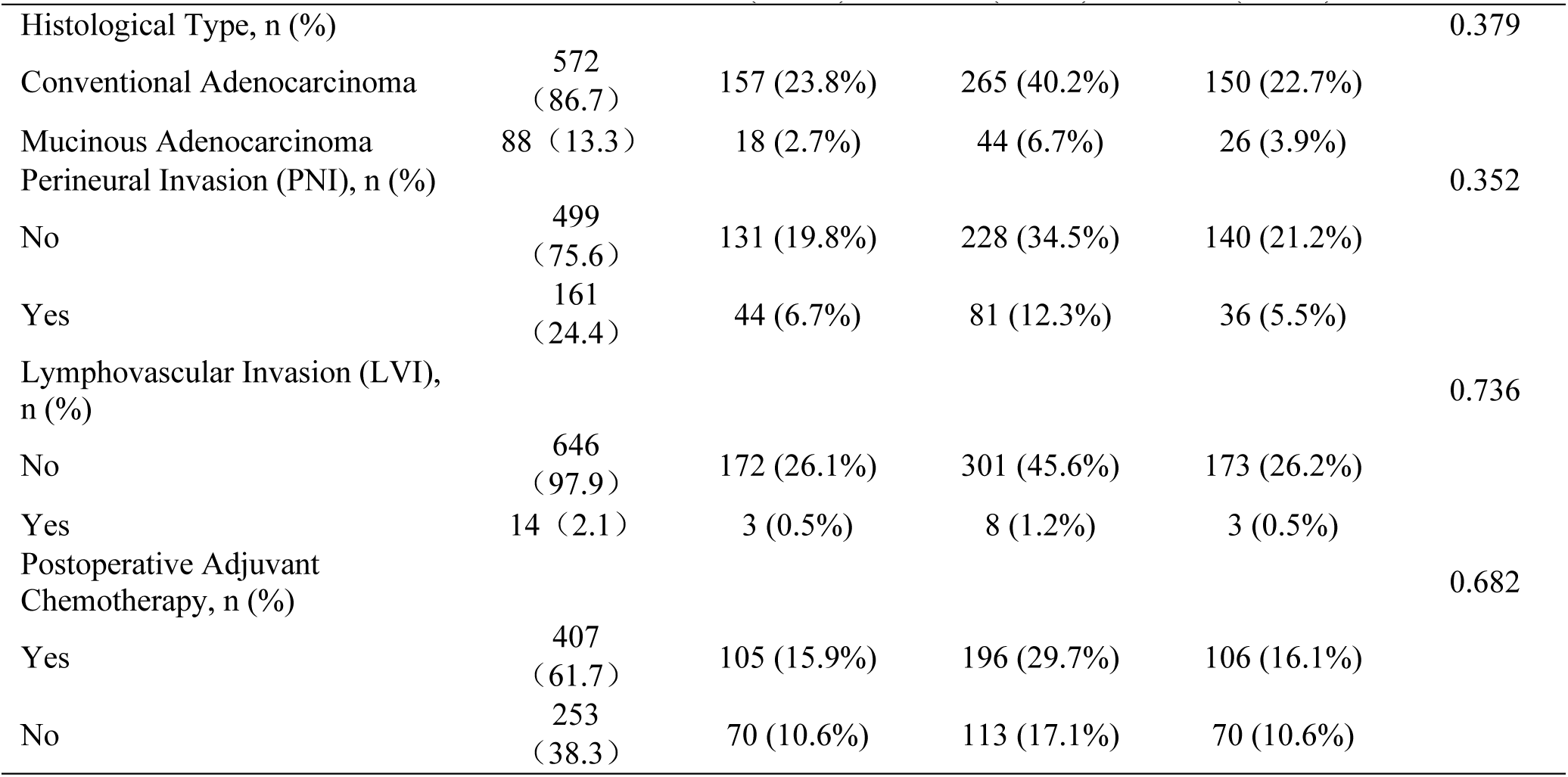
Baseline clinical characteristics of CRC patients stratified by ANPG grade.

| Characteristic | Total Cases<br>(n=660) | ANPG Grade<br>0<br>(n=175) | ANPG Grade<br>1<br>(n=309) | ANPG Grade<br>2<br>(n=176) | P-value |
| --- | --- | --- | --- | --- | --- |
| Age<br>[years, M (P25, P75)] | 61 (53, 68) | 60 (52, 65) | 61 (54, 68) | 64 (55.75, 70) | 0.001 |
| Gender, n (%) |  |  |  |  | 0.349 |
| Male | 353<br>(53.5) | 86 (13%) | 173 (26.2%) | 94 (14.2%) |  |
| Female | 307<br>(46.5) | 89 (13.5%) | 136 (20.6%) | 82 (12.4%) |  |
| TNM Stage, n (%) |  |  |  |  | 0.200 |
| Stage I | 102<br>(15.5) | 38 (5.8%) | 44 (6.7%) | 20 (3%) |  |
| Stage II | 298<br>(45.2) | 76 (11.5%) | 140 (21.2%) | 82 (12.4%) |  |
| Stage III | 249<br>(37.7) | 59 (8.9%) | 120 (18.2%) | 70 (10.6%) |  |
| Stage IVA | 11 (1.7) | 2 (0.3%) | 5 (0.8%) | 4 (0.6%) |  |
| Histological Type, n (%) |  |  |  |  | 0.379 |
| Conventional Adenocarcinoma | 572<br>(86.7) | 157 (23.8%) | 265 (40.2%) | 150 (22.7%) |  |
| Mucinous Adenocarcinoma | 88 (13.3) | 18 (2.7%) | 44 (6.7%) | 26 (3.9%) |  |
| Perineural Invasion (PNI), n (%) |  |  |  |  | 0.352 |
| No | 499<br>(75.6) | 131 (19.8%) | 228 (34.5%) | 140 (21.2%) |  |
| Yes | 161<br>(24.4) | 44 (6.7%) | 81 (12.3%) | 36 (5.5%) |  |
| Lymphovascular Invasion (LVI),<br>n (%) |  |  |  |  | 0.736 |
| No | 646<br>(97.9) | 172 (26.1%) | 301 (45.6%) | 173 (26.2%) |  |
| Yes | 14 (2.1) | 3 (0.5%) | 8 (1.2%) | 3 (0.5%) |  |
| Postoperative Adjuvant<br>Chemotherapy, n (%) |  |  |  |  | 0.682 |
| Yes | 407<br>(61.7) | 105 (15.9%) | 196 (29.7%) | 106 (16.1%) |  |
| No | 253<br>(38.3) | 70 (10.6%) | 113 (17.1%) | 70 (10.6%) |  |

As of the final follow-up date (December 16, 2024), the median follow-up duration was 2442 days (IQR: 2117–2537). During this period, 108 tumor-related deaths were recorded, resulting in an overall censoring rate of 83.6%.

### ANPG Distribution and Baseline Characteristics

According to the ANPG classification, the cohort included 175 patients (26.5%) classified as grade 0, 309 (46.8%) as grade 1, and 176 (26.7%) as grade 2. Baseline characteristics stratified by ANPG grade are shown in Table 1. Patients with higher ANPG grades were significantly older (P = 0.001). No significant differences were observed across the three groups in terms of sex, TNM stage, histological type, perineural invasion, lymphovascular invasion, or receipt of adjuvant chemotherapy (all P > 0.05).

### Discriminative Performance of ANPG Compared to Other Biomarkers

Receiver operating characteristic (ROC) curve analysis was performed to evaluate the discriminative ability of ANPG and other inflammatory/nutritional markers in predicting overall survival (OS) (Table 2, Fig 1). Among all assessed biomarkers, ANPG exhibited the highest area under the curve (AUC) value (0.637, 95% CI: 0.588–0.687, P < 0.001). DeLong’s test demonstrated that the AUC of ANPG was significantly greater than those of the neutrophil-to-lymphocyte ratio (NLR) (AUC = 0.574, P = 0.047), platelet-to-lymphocyte ratio (PLR) (AUC = 0.519, P = 0.002), systemic immune-inflammation index (SII) (AUC = 0.537, P = 0.001), and F-NLR score (AUC = 0.591, P = 0.049). For the fibrinogen-to-albumin ratio (FAR), the continuous variable (FAR_cont, log-transformed using the natural logarithm (Ln) to account for non-normal distribution), yielded an AUC of 0.596, whereas the categorical variable (FAR_cat) resulted in an AUC of 0.589 (95% CI: 0.540–0.639, P = 0.003). Additionally, ANPG showed superior predictive performance compared to albumin (AUC = 0.615) and neutrophil count (AUC = 0.560) as individual parameters.

**Fig 1.**
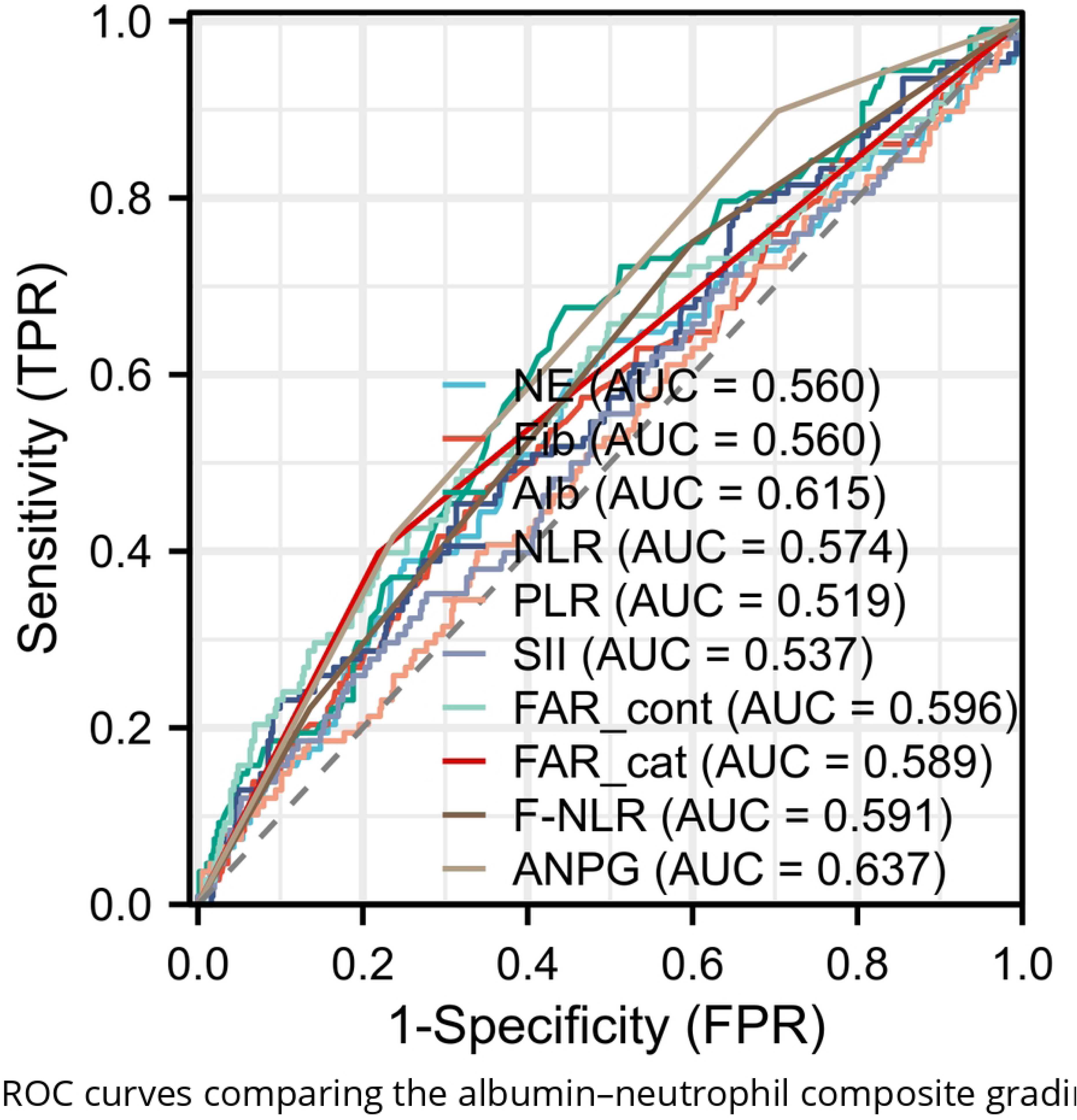
ROC curves comparing the albumin–neutrophil composite grading (ANPG) system with other inflammatory/nutritional biomarkers for predicting overall survival (OS) in colorectal cancer (CRC) patients. ANPG achieved the highest AUC (0.637) and was significantly superior to NLR (0.574), PLR (0.519), SII (0.537), FAR_cont(0.596), FAR_cat(0.589)and F-NLR (0.591); DeLong test, all P < 0.05.

**Table 2:** Results of ROC curve analysis for prognostic indicators in CRC patients.

| Indicator | AUC | Confidence Interval (CI) | Sensitivity (%) | Specificity (%) | Youden Index | Cut-off Value | P-value |
| --- | --- | --- | --- | --- | --- | --- | --- |
| NE | 0.560 | 0.498 – 0.622 | 0.63889 | 0.50543 | 0.14432 | 3.755 | 0.049 |
| Fib | 0.560 | 0.499 – 0.621 | 0.41667 | 0.70833 | 0.125 | 3.595 | 0.049 |
| Alb | 0.615 | 0.558 – 0.673 | 0.67593 | 0.55435 | 0.23027 | 42.35 | <0.001 |
| NLR | 0.574 | 0.514 – 0.634 | 0.4537 | 0.68659 | 0.1403 | 3.0157 | 0.015 |
| PLR | 0.519 | 0.458 – 0.580 | 0.71296 | 0.34783 | 0.060789 | 145.43 | 0.524 |
| SII | 0.537 | 0.476 – 0.598 | 0.75 | 0.3279 | 0.077899 | 492.02 | 0.220 |
| FAR_cont | 0.596 | 0.534–0.659 | 0.39815 | 0.7808 | 0.17895 | 0.091734 | 0.002 |
| FAR_cat | 0.589 | 0.540–0.639 | 0.39815 | 0.7808 | 0.17895 | 0.5 | 0.003 |
| F-NLR | 0.591 | 0.538 – 0.644 | 0.75 | 0.40036 | 0.15036 | 0.5 | 0.003 |
| ANPG | 0.637 | 0.588 – 0.687 | 0.89815 | 0.2971 | 0.19525 | 0.5 | <0.001 |

Abbreviations: NE, Neutrophils; Alb, Albumin; NLR, Neutrophil-to-Lymphocyte Ratio; PLR, Platelet-to-Lymphocyte Ratio; SII, Systemic Immune-Inflammation Index; FAR\_cont, Fibrinogen-to-albumin ratio (FAR) Continuous variable after natural logarithm (Ln) transformation; FAR\_cat, Fibrinogen-to-albumin ratio (FAR) Categorical; F-NLR, Fibrinogen-Neutrophil-Lymphocyte Ratio; ANPG, Albumin-Neutrophil Combined Prognostic Grade

### ANPG and Overall Survival

Kaplan-Meier survival analysis revealed significant differences in overall survival (OS) across the three ANPG grade groups (log-rank χ² = 24.423, P < 0.001) (Fig 2). Mean OS durations were 2442.743 ± 31.217 days for grade 0, 2296.343 ± 33.854 days for grade 1, and 2116.641 ± 59.011 days for grade 2. The corresponding 5-year OS rates were 93.7%, 83.2%, and 74.4%, respectively. Pairwise comparisons showed statistically significant differences between all ANPG grade groups (all P < 0.05).

**Fig 2.**
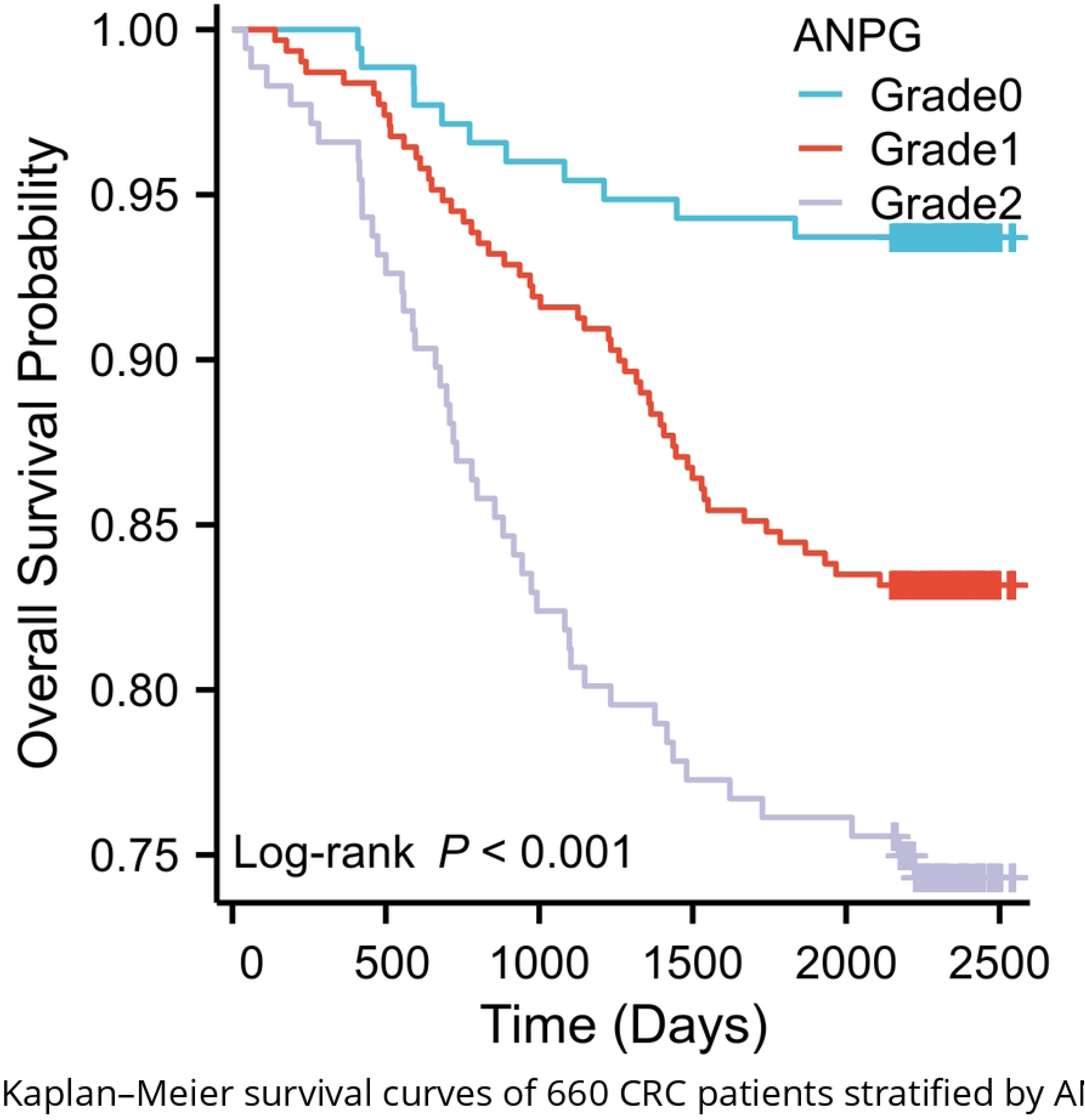
Kaplan–Meier survival curves of 660 CRC patients stratified by ANPG grade. Five-year OS rates: Grade 0, 93.7 % (n = 175); Grade 1, 83.2 % (n = 309); Grade 2, 74.4 % (n = 176). Higher grade was associated with poorer OS (log-rank χ² = 24.423, P < 0.001; all pairwise comparisons P < 0.05).

### Univariate and Multivariate Cox Regression Analysis

Univariate Cox regression analysis identified several factors significantly associated with overall survival (OS) (Table 3), including age, carcinoembryonic antigen (CEA), carbohydrate antigen 19-9 (CA19-9), systemic immune-inflammation index (SII), fibrinogen-to-neutrophil-lymphocyte ratio (F-NLR), albumin-to-neutrophil-lymphocyte ratio prognostic grade (ANPG), TNM stage, histological type, and postoperative adjuvant chemotherapy (all P < 0.10).

**Table 3:** Univariate Cox regression analysis of OS in CRC patients.

| Factor | Hazard Ratio<br>(95% Confidence<br>Interval) | P-value |
| --- | --- | --- |
| Age (per 1-year increase) | 1.033 (1.014 - 1.053) | < 0.001 |
| Gender (Male vs Female) | 0.891 (0.611 - 1.299) | 0.548 |
| TNM Stage |  | <0.001 |
| Stage II vs Stage I | 2.705 (0.812 - 9.008) | 0.105 |
| Stage III vs Stage I | 12.185 (3.842 - 38.644) | < 0.001 |
| Stage IVA vs Stage I | 31.848 (8.224 - 123.324) | < 0.001 |
| Histological Type (Mucinous adenocarcinoma vs Conventional adenocarcinoma) | 2.170 (1.387 - 3.394) | < 0.001 |
| Perineural Invasion (Yes vs No) | 1.405 (0.933 - 2.116) | 0.103 |
| Lymphovascular Invasion (Yes vs No) | 1.728 (0.636 - 4.690) | 0.283 |
| Postoperative Adjuvant Chemotherapy (Yes vs No) | 1.843 (1.198 - 2.834) | 0.005 |
| CEA (per 1 ng/mL increase) | 1.009 (1.006 - 1.012) | < 0.001 |
| CA19-9 (per 1 U/mL increase) | 1.003 (1.002 - 1.004) | < 0.001 |
| CA724 | 1.004 (0.998 - 1.011) | 0.172 |
| Neutrophil-to-Lymphocyte Ratio (NLR) | 1.055 (0.989 - 1.126) | 0.105 |
| Platelet-to-Lymphocyte Ratio (PLR) | 1.001 (0.999 - 1.003) | 0.244 |
| Systemic Immune-Inflammation Index (SII) | 1.000 (1.000 - 1.000) | 0.064 |
| Fibrinogen-to-albumin ratio (FAR)_cont | 3.564 (1.696 - 7.489) | < 0.001 |
| Fibrinogen-to-albumin ratio (FAR)_cat | 2.201 (1.497 - 3.235) | < 0.001 |
| Fibrinogen-to-Neutrophil Lymphocyte Ratio (F-NLR) (1 vs 0) | 1.758 (1.112 - 2.779) | 0.016 |
| Fibrinogen-to-Neutrophil Lymphocyte Ratio (F-NLR) (2 vs 0) | 2.441 (1.408 - 4.230) | 0.001 |
| ANPG (1 vs 0) | 2.797 (1.460 - 5.361) | 0.002 |
| ANPG (2 vs 0) | 4.593 (2.376 - 8.881) | < 0.001 |

Notably, both the continuous (FAR_cont; HR = 3.564, 95% CI: 1.696–7.489, P < 0.001) and categorical (FAR_cat; HR = 2.201, 95% CI: 1.497–3.235, P < 0.001) forms of the fibrinogen-to-albumin ratio (FAR) were significantly associated with OS in univariate analysis. To minimize multicollinearity due to inclusion of both FAR variants and to enhance clinical applicability, only the categorical FAR variable (FAR_cat) was included in the multivariate Cox proportional hazards regression model. However, after adjustment for ANPG, TNM stage, age, CA19-9, CEA, and histological type, FAR_cat no longer retained statistical significance (all P ≥ 0.53). This suggests that when ANPG—a more robust composite inflammatory-nutritional marker—was included in the model, FAR did not independently predict OS in this patient cohort.

Multivariate Cox proportional hazards regression analysis (Table 4) identified several independent prognostic factors (all variance inflation factors <4, indicating acceptable multicollinearity): ANPG (grade 1 vs. 0: HR = 2.190, 95% CI: 1.131–4.240, P = 0.020; grade 2 vs. 0: HR = 3.256, 95% CI: 1.576–6.727, P < 0.001), age (per 1-year increase: HR = 1.032, 95% CI: 1.013–1.052, P < 0.001), CA19-9 (per 1 U/mL increase: HR = 1.002, 95% CI: 1.001–1.004, P = 0.003), histological subtype (mucinous adenocarcinoma vs. conventional adenocarcinoma: HR = 1.954, 95% CI: 1.222–3.124, P = 0.005), and TNM stage (stage III vs. I: HR = 8.655, 95% CI: 2.563–29.222, P < 0.001; stage IVA vs. I: HR = 22.896, 95% CI: 5.344–98.102, P < 0.001) were identified as independent prognostic factors. Although CEA did not achieve statistical significance in the final multivariate model (P = 0.075), it was retained due to its established clinical relevance and borderline significant association in univariate analysis (HR = 1.009, 95% CI: 1.006–1.012, P < 0.001). CEA was incorporated as a continuous variable in the nomogram, as preliminary analyses failed to identify an optimal cutoff with adequate discriminative performance (AUC = 0.582, P = 0.012).

**Table 4:** Multivariate Cox regression analysis of OS in CRC patients.

| Factor | Hazard Ratio<br>(95% Confidence<br>Interval) | P-value | VIF |
| --- | --- | --- | --- |
| Age (per 1-year increase) | 1.032 (1.013~1.052) | <0.001 | 1.084 |
| TNM Stage |  | <0.001 | 1.373 |
| Stage II vs Stage I | 1.837 (0.522 - 6.466) | 0.343 | — |
| Stage III vs Stage I | 8.655 (2.563 - 29.222) | < 0.001 | — |
| Stage IVA vs Stage I | 22.896 (5.344 - 98.102) | < 0.001 | — |
| Histological Type (Mucinous vs Conventional Adenocarcinoma) | 1.954 (1.222 - 3.124) | 0.005 | 1.039 |
| Postoperative Adjuvant Chemotherapy (Yes vs No) | 1.120 (0.696 - 1.803) | 0.641 | 1.381 |
| CEA (per 1 ng/mL increase) | 1.004 (1.000 - 1.008) | 0.075 | 1.236 |
| CA19-9 (per 1 U/mL increase) | 1.002 (1.001~1.004) | 0.003 | 1.221 |
| Systemic Immune-Inflammation Index (SII) | 1.000 (0.999 - 1.000) | 0.131 | 1.475 |
| FAR_cat | 1.164 (0.721 - 1.879) | 0.534 | 1.439 |
| Fibrinogen-to-Neutrophil Lymphocyte Ratio (F-NLR) |  |  | 1.706 |
| 1 vs 0 | 1.266 (0.771 - 2.080) | 0.352 | — |
| 2 vs 0 | 1.513 (0.714 - 3.208) | 0.280 | — |
| ANPG |  |  | 1.399 |
| 1 vs 0 | 2.190 (1.131 - 4.240) | 0.020 | — |
| 2 vs 0 | 3.256 (1.576 - 6.727) | 0.001 | — |

Abbreviations: VIF, Variance Inflation Factor

The proportional hazards assumption was satisfied for all covariates in the multivariate model (global test: χ² = 18.941, P = 0.167).

### Forest Plot Analysis

A forest plot was constructed to visualize the effect sizes of the independent prognostic factors (Fig 3). The results revealed a hierarchical risk stratification, with TNM stage IVA conferring the highest mortality risk (HR = 22.896), followed by stage III (HR = 8.655). ANPG exhibited a graded increase in risk from grade 0 to grade 2, while age, CA19-9, and histological subtype contributed additional prognostic discrimination.

**Fig 3.**
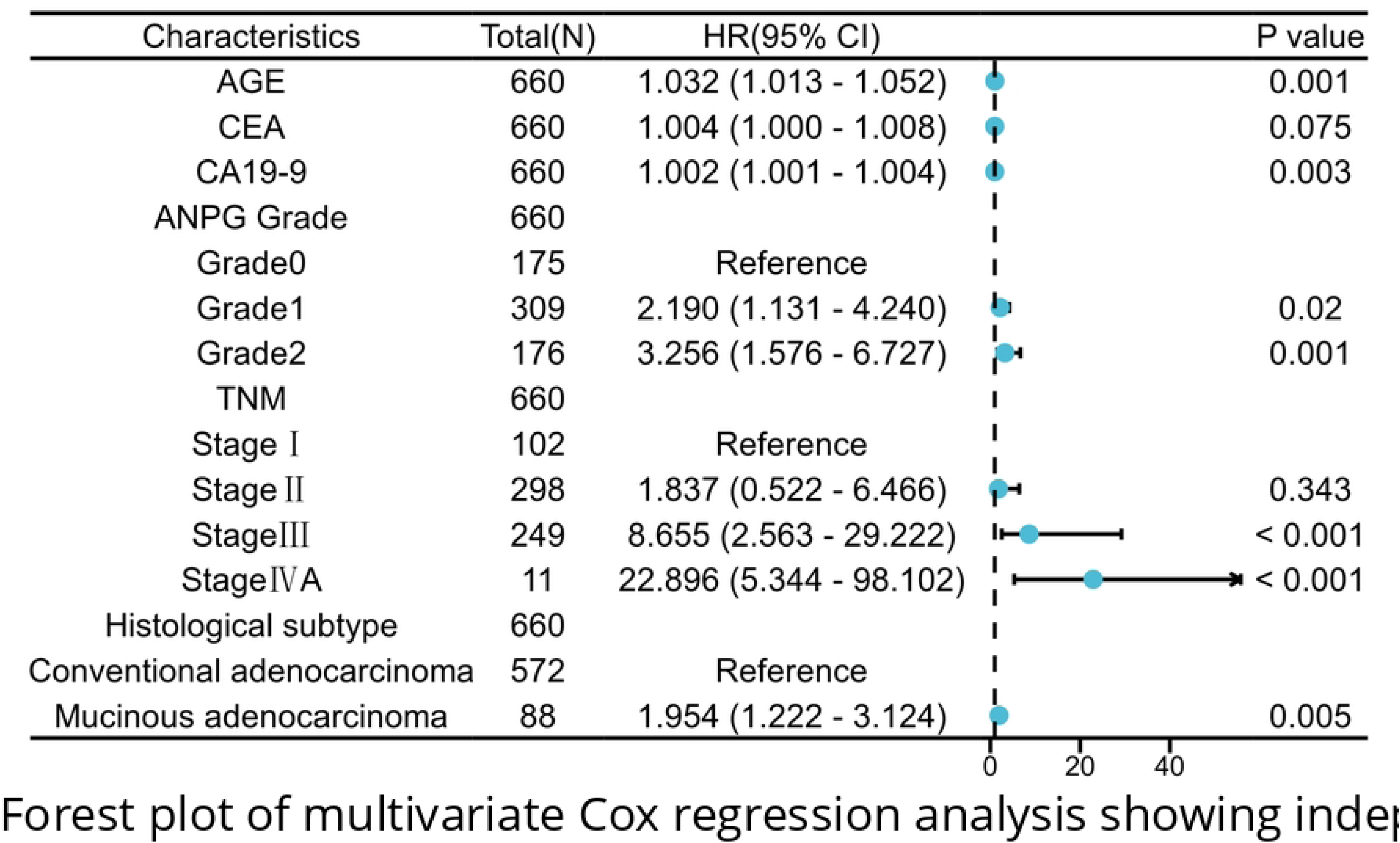
Forest plot of multivariate Cox regression analysis showing independent prognostic factors in CRC. Key hazard ratios (95 % CI): TNM stage IVA vs I, 22.896 (5.344–98.102); ANPG Grade 2 vs 0, 3.256 (1.576–6.727); age (per 1-year increase), 1.032 (1.013–1.052)

### Nomogram Development and Validation

#### 1. Construction of the Nomogram Model and Variable Selection

A nomogram was developed to predict 1-year, 3-year, and 5-year overall survival (OS) probabilities in patients with colorectal cancer, incorporating six key variables: ANPG, age, CA19-9, CEA, histological type, and TNM stage (Fig 4). Univariate Cox regression analysis demonstrated that CEA was significantly associated with OS (HR = 1.009, 95% CI: 1.006–1.012, P < 0.001). Model comparison showed that inclusion of CEA improved the concordance index (C-index) to 0.806 (95% CI: 0.788–0.824), compared to 0.800 (95% CI: 0.782–0.818) when excluded. Therefore, CEA was retained in the final model.

**Fig 4.**
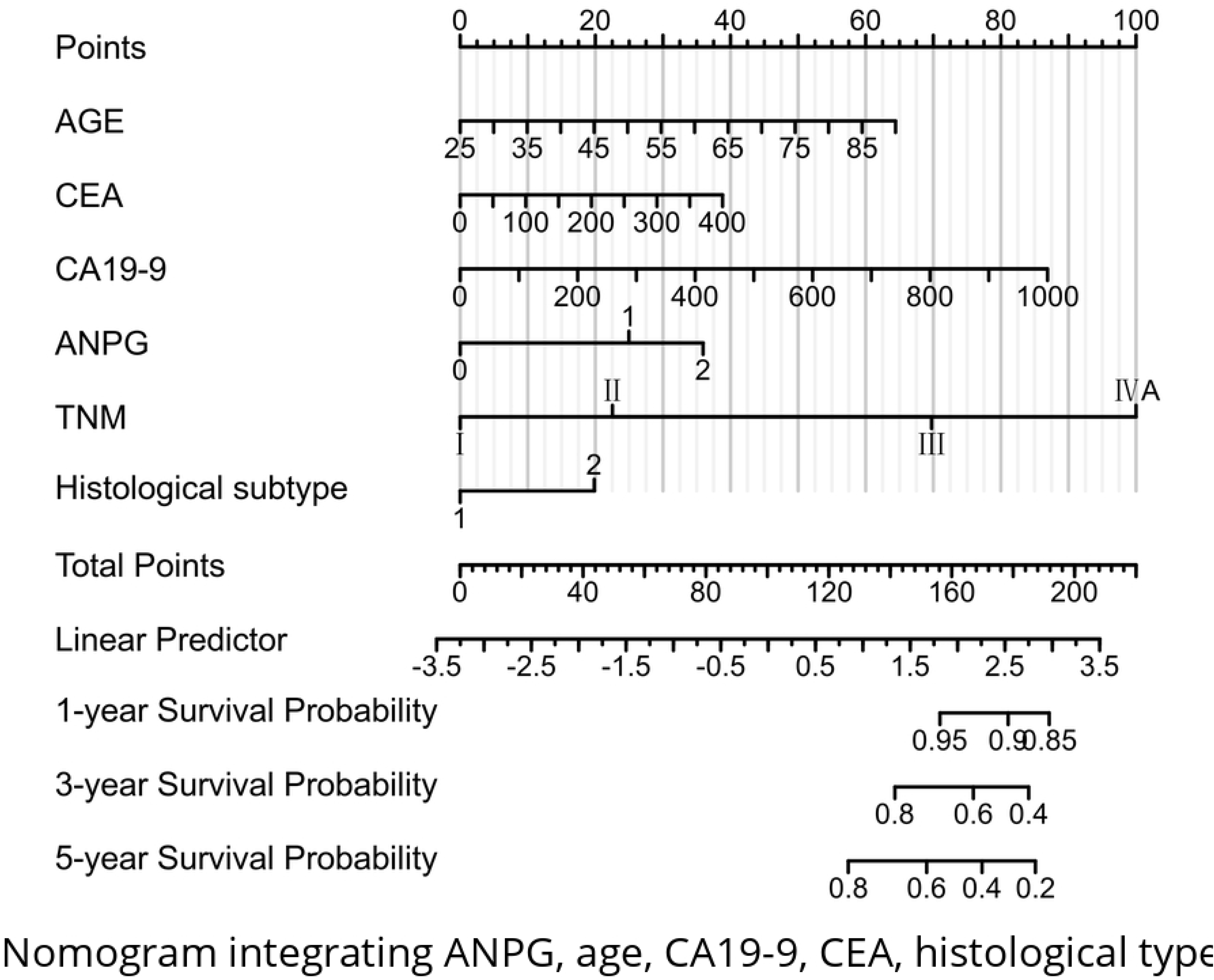
Nomogram integrating ANPG, age, CA19-9, CEA, histological type and TNM stage to predict 1-, 3-and 5-year OS after radical resection in CRC patients. Model discrimination: C-index = 0.806 (95 % CI 0.788–0.824).

#### 2. Validation of Model Performance

The nomogram demonstrated strong discriminative ability, with a C-index of 0.806 (95% CI: 0.788–0.824). Calibration plots showed excellent agreement between predicted and observed 1-year, 3-year, and 5-year OS probabilities (Fig 5). Internal validation using 800 bootstrap resampling iterations (100 samples per iteration) confirmed the robustness and stability of the model.

**Fig 5.**
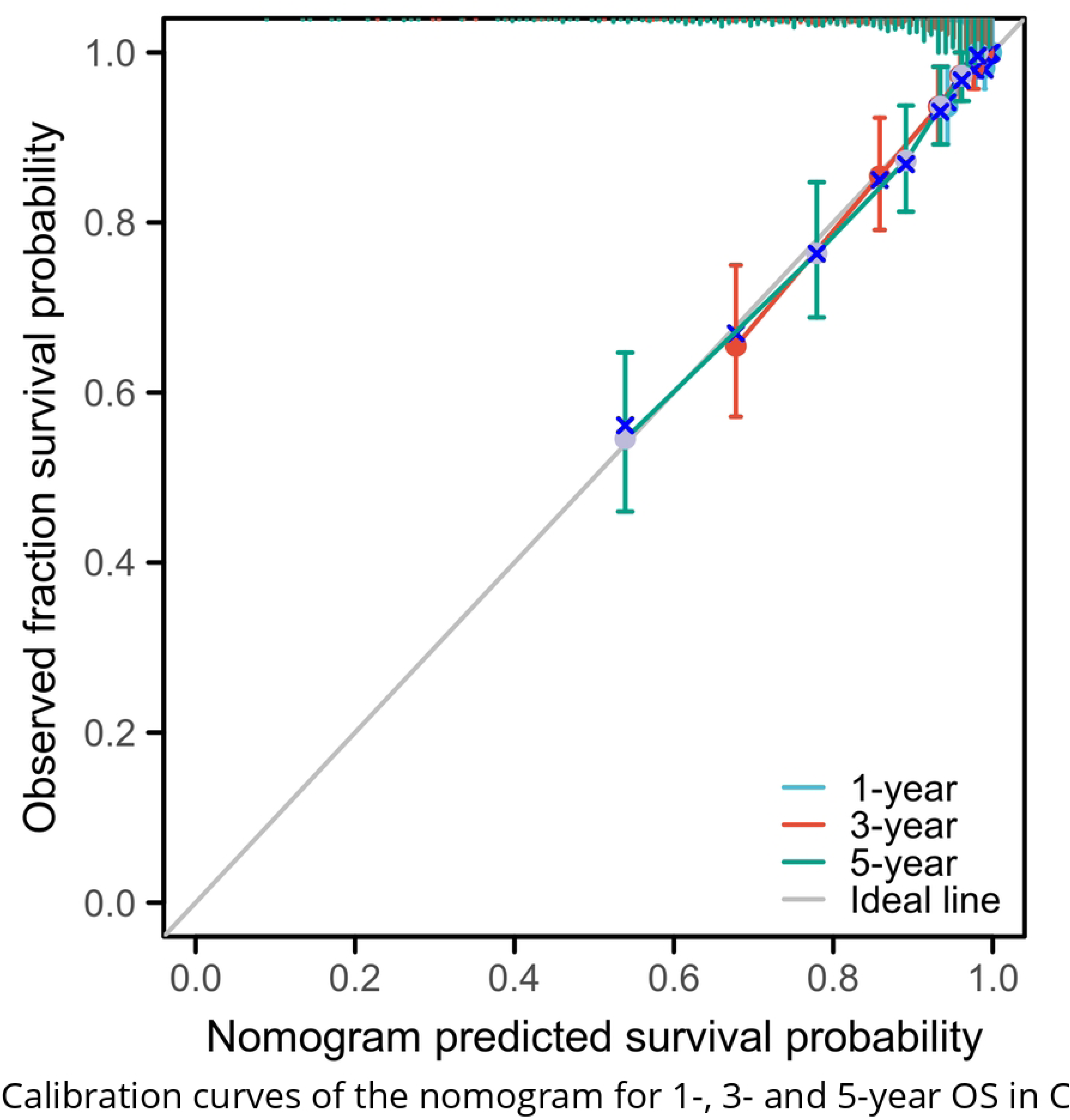
Calibration curves of the nomogram for 1-, 3– and 5-year OS in CRC patients. Bootstrap validation (800 resamples, 100 cases per iteration) demonstrated good agreement between predicted and observed survival probabilities.

#### 3. Time-Dependent Receiver Operating Characteristic (ROC) Analysis

Time-dependent ROC analysis was performed to evaluate the dynamic predictive accuracy of the nomogram. The area under the curve (AUC) values for 1-year, 3-year, and 5-year OS predictions were 0.841, 0.849, and 0.837, respectively, all significantly higher than those of the conventional TNM staging system (AUCs: 0.646, 0.759, and 0.749; all P < 0.05) (Fig 6, Fig 7, Fig 8).

**Fig 6.**
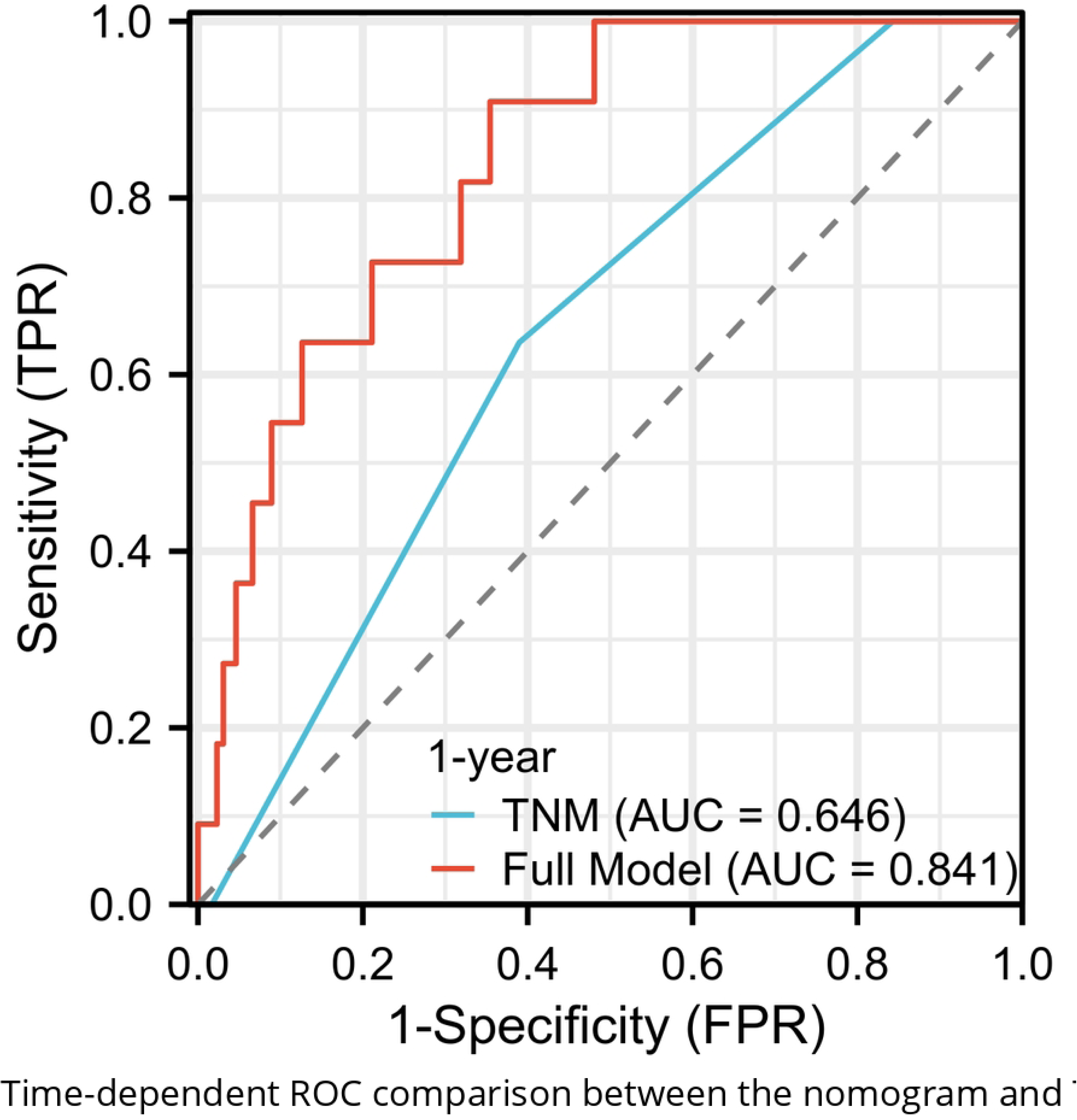
Time-dependent ROC comparison between the nomogram and TNM staging for 1-year OS in CRC patients: nomogram AUC = 0.841 vs TNM 0.646, P < 0.05.

**Fig 7.**
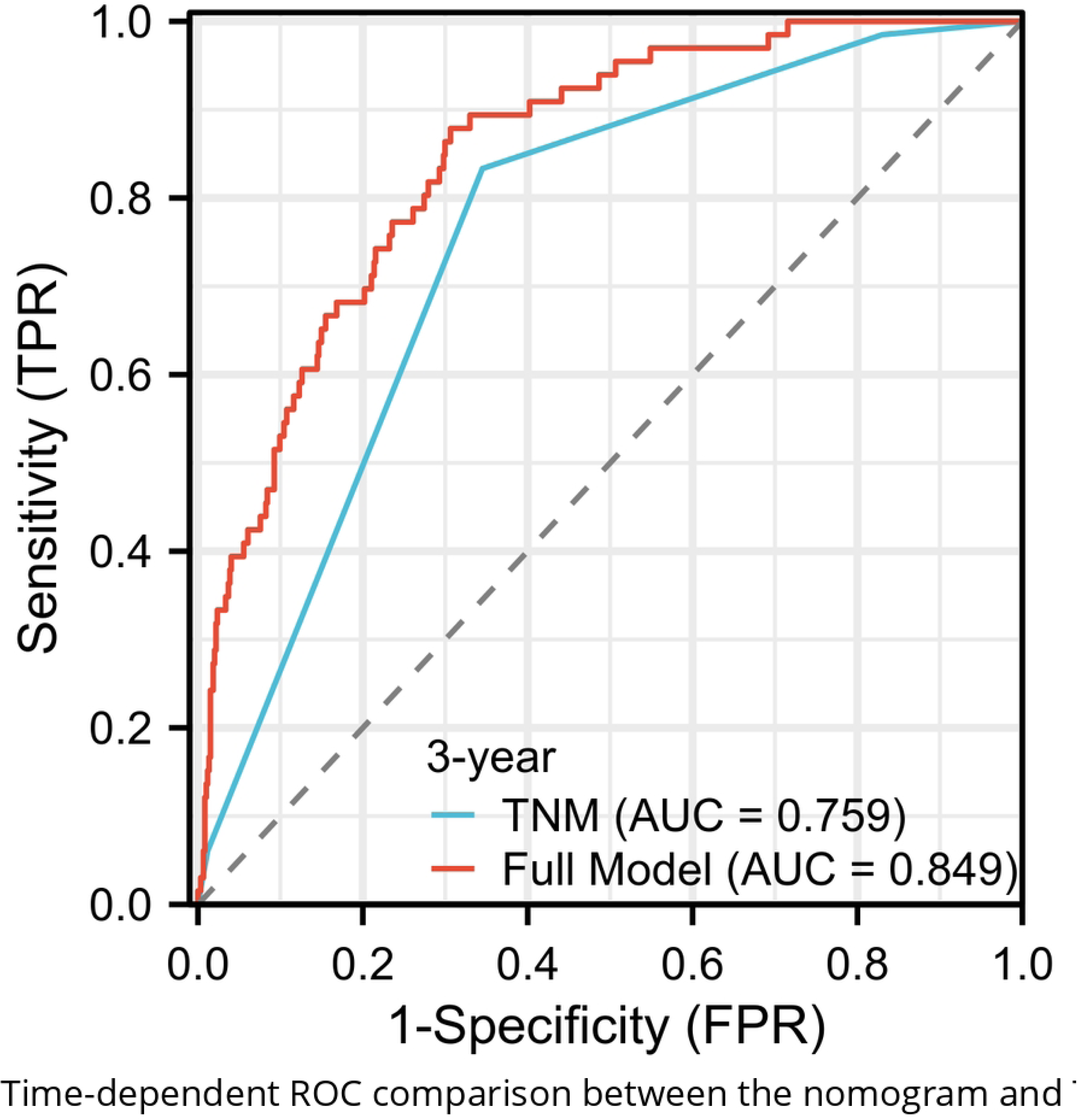
Time-dependent ROC comparison between the nomogram and TNM staging for 3-year OS in CRC patients: nomogram AUC = 0.849 vs TNM 0.759, P < 0.05.

**Fig 8.**
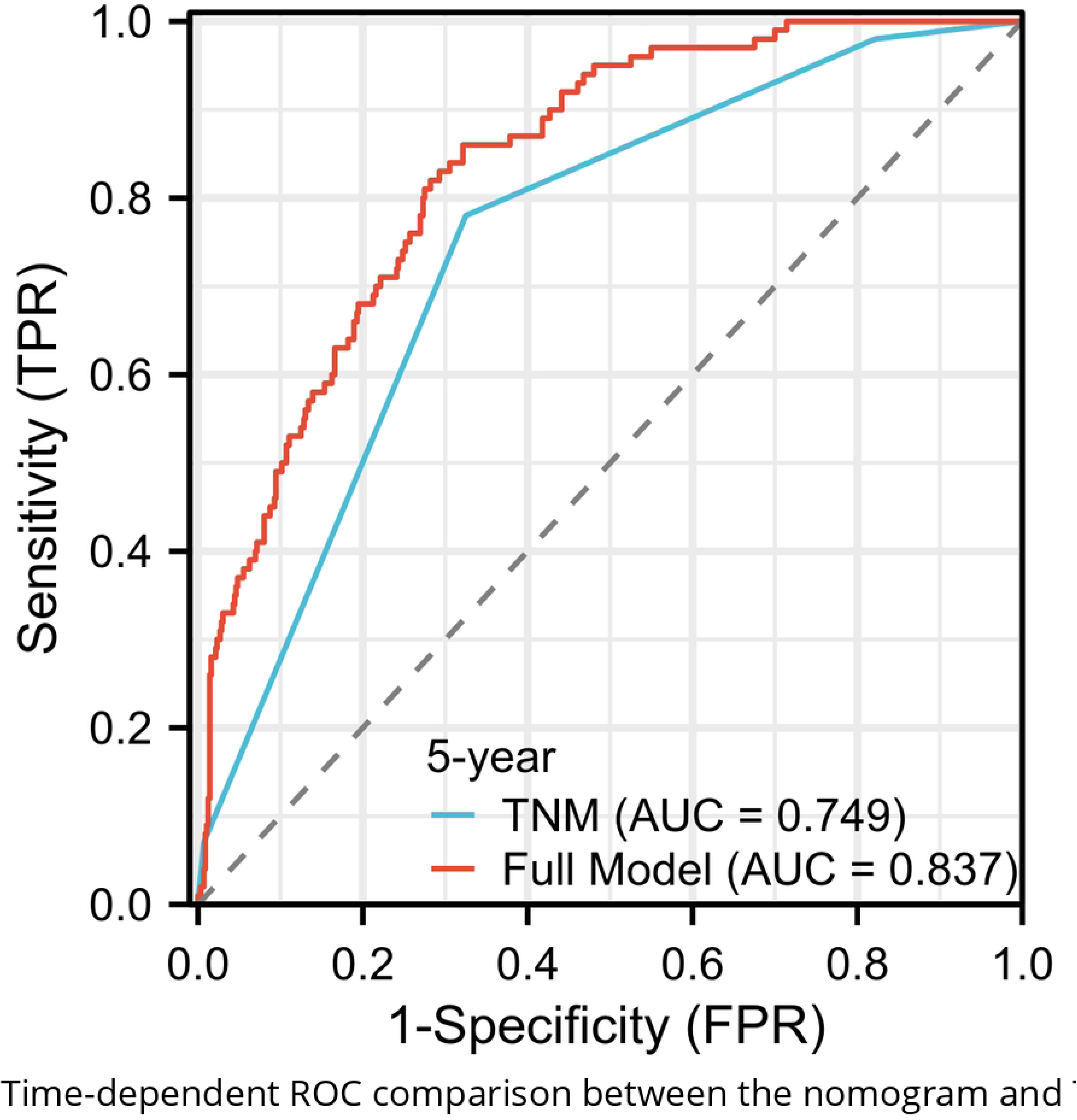
Time-dependent ROC comparison between the nomogram and TNM staging for 5-year OS in CRC patients: nomogram AUC = 0.837 vs TNM 0.749, P < 0.05.

#### 4. Decision Curve Analysis (DCA)

To assess clinical utility, decision curve analysis (DCA) was conducted to quantify the net benefit of the nomogram for predicting 5-year OS across a range of threshold probabilities (0–1.0) (Fig 9). Within the clinically meaningful range (0.05–0.70), the nomogram consistently provided greater net benefit than TNM staging alone. Beyond 0.70, the net benefits declined for both strategies, though the nomogram remained superior. These findings support the enhanced clinical applicability of the proposed nomogram over traditional TNM staging.

**Fig 9.**
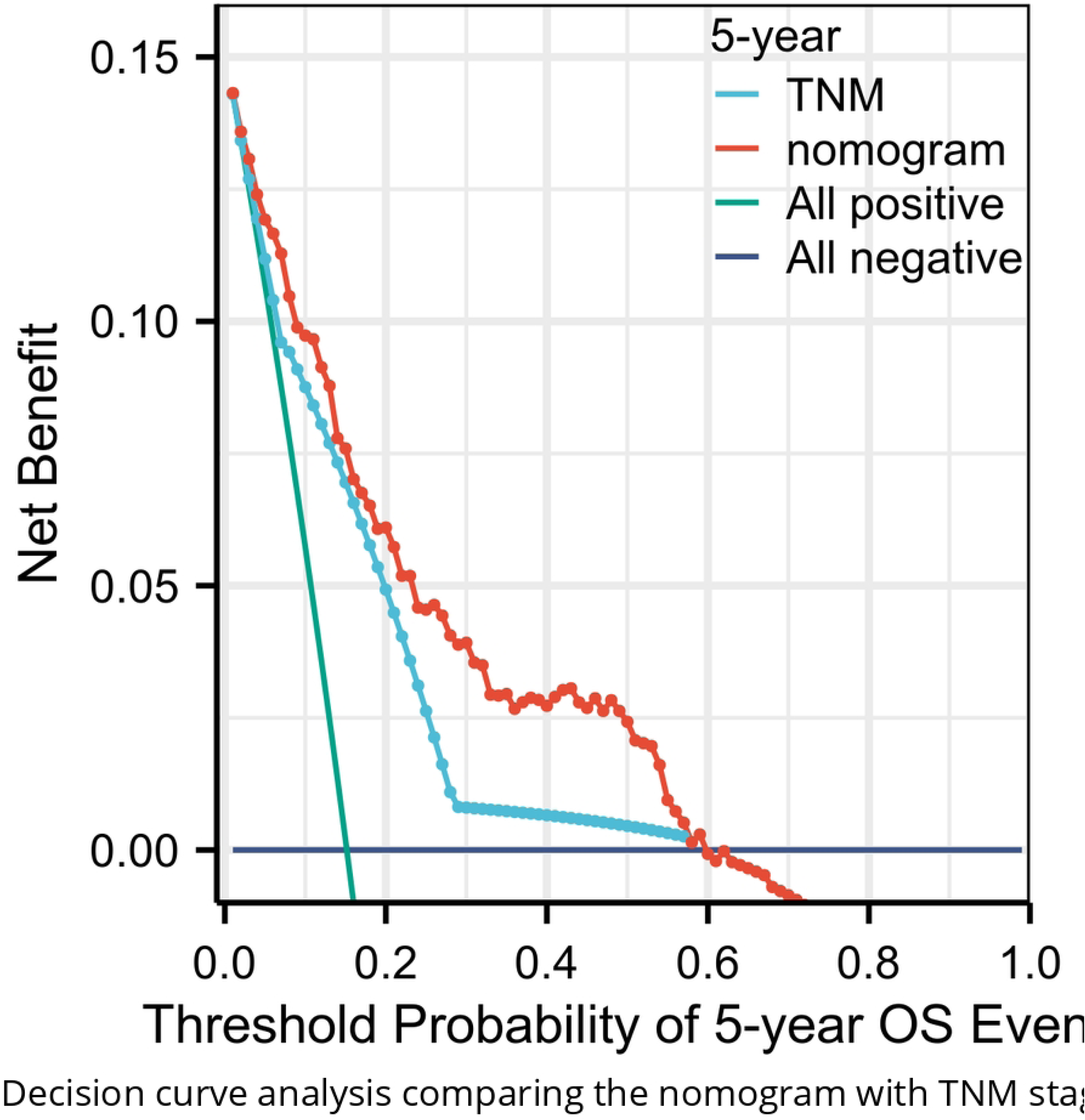
Decision curve analysis comparing the nomogram with TNM staging and default strategies (treat-all/treat-none). The nomogram yielded higher net benefit across a wide range of threshold probabilities, indicating superior clinical utility.

## Discussion

This study establishes the preoperative albumin-neutrophil prognostic grade (ANPG) system as an independent and robust prognostic indicator for patients with colorectal cancer (CRC) undergoing radical resection. Our results demonstrate that ANPG exhibits superior discriminative ability compared to conventional inflammatory and nutritional biomarkers, and its integration into a nomogram significantly enhances individualized survival prediction beyond the traditional TNM staging system.

The prognostic value of ANPG in CRC is rooted in the pathophysiological interplay between nutritional status and systemic inflammation in cancer progression., As a negative acute-phase reactant, albumin serves as a dual biomarker reflecting both nutritional reserve and systemic inflammatory response. Hypoalbuminemia is associated with impaired immune function, increased risk of infection, and reduced tolerance to therapy^[19, 24]^. Concurrently, neutrophilia reflects an exaggerated systemic inflammatory response, which promotes tumor progression through multiple mechanisms, including angiogenesis induction, extracellular matrix remodeling, and suppression of antitumor immunity^[21, 22]^. By integrating these two clinically relevant parameters, the ANPG system provides a comprehensive assessment of the host inflammatory-nutritional axis, demonstrating greater prognostic accuracy than individual markers or other composite scores.

Our findings corroborate and extend previous evidence from studies on other malignancies. Sun et al. first reported the prognostic utility of the albumin-neutrophil combined grade in non-small cell lung cancer, showing improved predictive performance over single parameters^[23]^. Wang et al. showed that the albumin-neutrophil-to-lymphocyte ratio could predict survival and postoperative complications in elderly patients with CRC^[25]^. The current study builds on these observations by confirming that ANPG serves as an independent predictor of clinical outcomes in a large CRC cohort, outperforming multiple established inflammatory-nutritional indices.

Comparative analysis revealed that ANPG exhibited significantly superior discriminative capability (AUC=0.637) relative to the neutrophil-to-lymphocyte ratio (NLR, 0.574), platelet-to-lymphocyte ratio (PLR, 0.519), systemic immune-inflammation index (SII, 0.537), and fibrinogen-NLR (F-NLR, 0.591) (all P<0.05 by DeLong’s test). Notably, ANPG incorporates only two routinely measured preoperative parameters—serum albumin and neutrophil counts—offering distinct clinical utility without necessitating additional diagnostic expenditures. Emerging evidence from randomized controlled trials suggests that perioperative nutritional intervention may mitigate postoperative complications in patients with elevated inflammatory markers^[26]^. Thus, ANPG represents a practical clinical tool for identifying high-risk patients who may benefit from intensive preoperative nutritional optimization. Specifically,, patients classified as ANPG grade 2 (exhibiting concurrent hypoalbuminemia and neutrophilia) might experience reduced complication rates following targeted nutritional support, though this hypothesis requires prospective validation.

In contrast to composite scoring systems that rely on multiple biomarkers, ANPG offers enhanced operational simplicity and greater feasibility in resource-constrained settings. Its prognostic superiority likely stems from the simultaneous evaluation of nutritional status and systemic inflammation, thereby providing a more comprehensive assessment of host-tumor dynamics. This streamlined methodology renders ANPG particularly suitable for implementation in primary care facilities or institutions with limited diagnostic infrastructure, facilitating broader clinical applicability.

Multivariate analysis yields particularly informative results. Although the fibrinogen-to-albumin ratio (FAR) showed significant prognostic association in univariate analysis, this significance was lost after adjustment for ANPG and other covariates, suggesting that ANPG captures more comprehensive prognostic information than FAR. This finding contrasts with some prior reports^[27, 28]^, but aligns with a recent meta-analysis indicating inconsistent prognostic value of FAR in colorectal cancer^[16]^. The independent prognostic significance of ANPG, in addition to well-established clinical factors such as age, CA19-9, histological subtype, CEA, and TNM stage, underscores its incremental value in risk stratification.

Carcinoembryonic antigen (CEA) is a well-established tumor marker in the management of colorectal cancer, with levels correlating with tumor burden and risk of recurrence^[29]^. Our comparative analysis demonstrated that incorporation of CEA improved the nomogram’s discriminative accuracy, increasing the concordance index (C-index) from 0.800 to 0.806, thus confirming its incremental prognostic value. This finding aligns with existing evidence indicating that conventional tumor markers can complement inflammatory and nutritional biomarkers to enhance predictive model performance^[30, 31]^. Although CEA did not reach statistical significance in multivariate analysis (P = 0.075), its inclusion in the model is justified by the observed improvement in predictive capability and its established clinical relevance. This approach underscores the principle that prognostic models should balance statistical rigor with clinical meaningfulness^[32]^.

The developed nomogram serves as a clinically applicable decision-support tool, demonstrating strong predictive performance with a concordance index (C-index) of 0.806. It exhibits superior calibration compared to existing prognostic nomograms for colorectal cancer that integrate TNM staging and inflammatory biomarkers^[33]^, offering more accurate individualized survival predictions than conventional staging systems alone. Time-dependent receiver operating characteristic (ROC) analysis confirmed enhanced discrimination, with consistently higher area under the curve (AUC) values at 1-, 3-, and 5-year follow-up timepoints. Decision curve analysis further validated its clinical utility, revealing greater net benefit across relevant threshold probabilities, supporting its potential to guide treatment decisions and optimize resource utilization. These results have significant clinical implications, as the nomogram may facilitate more precise identification of high-risk patients who could benefit from intensified surveillance, adjuvant therapy, or nutritional interventions.

Notably, patients with elevated ANPG scores were significantly older (P = 0.001). This association may be explained by age-related declines in nutritional status and increased systemic inflammation—both recognized contributors to tumor progression^[34, 35]^. Aging is linked to reduced albumin synthesis, dysregulated immune function, and chronic low-grade inflammation (termed inflammaging), which may collectively lead to higher ANPG scores and poorer clinical outcomes in older individuals.

This study has several key strengths. First, the relatively large sample size (n = 660) and extended follow-up duration (median 2442 days) provide robust statistical power and reliable survival estimates. Second, strict inclusion criteria—limited to R0-resected cases and exclusion of patients with confounding conditions—help minimize bias. Third, systematic comparison with multiple established inflammatory and nutritional markers highlights the superior prognostic performance of the ANPG score. Fourth, comprehensive statistical validation methods, including bootstrap resampling and time-dependent ROC analysis, strengthen the reliability and generalizability of the findings.

Nevertheless, several limitations should be acknowledged. First, as a single-center retrospective study, the potential for selection bias and limited external validity remains, highlighting the need for multi-center validation. Second, the cut-off values for albumin and neutrophil counts were determined based on the specific cohort, and their generalizability to other populations requires further investigation. Third, the absence of assessment of dynamic changes in the albumin-neutrophil composite grading (ANPG) during the postoperative period may represent an unexplored prognostic dimension. Fourth, the relatively low event rate (n = 108) constrained the performance of detailed subgroup analyses and precluded independent validation of the nomogram. Finally, the biological mechanisms underlying the prognostic impact of ANPG warrant further elucidation through prospective studies integrating molecular and immunological profiling.

## Conclusion

In summary, this study establishes the preoperative albumin-neutrophil composite grading (ANPG) system as a robust and independent prognostic indicator in colorectal cancer (CRC) patients undergoing radical resection. The ANPG demonstrates superior discriminative ability compared with conventional inflammatory and nutritional biomarkers. The proposed nomogram incorporating ANPG provides a clinically applicable tool for accurate survival prediction and risk stratification, potentially aiding therapeutic decision-making in CRC management. Further validation through prospective, multicenter clinical trials is warranted to confirm these findings and clarify the pathophysiological mechanisms underlying the prognostic value of ANPG.

## Data Availability

All relevant data are within the manuscript and its Supporting Information files.

## Acknowledgments

The authors acknowledge the Hebei Provincial Medical Science Research Project (No. 20240588) for administrative support and acknowledge Xiantao Academic Platform for technical assistance with statistical analysis and figure preparation.

## Author Contributions

Conceptualization: Xuelian Shi, Chaoxi Zhou, Chunfu Wan, Xiaoli Xu

Data curation: Guo Tian, Jiena Zhou, Haiyan Fu

Formal analysis: Xuelian Shi, Chunfu Wan

Investigation: Guo Tian, Jiena Zhou, Haiyan Fu

Methodology: Xuelian Shi, Chaoxi Zhou, Xiaoli Xu

Project administration: Xuelian Shi

Software: Guo Tian, Jiena Zhou

Supervision: Chaoxi Zhou, Chunfu Wan, Xiaoli Xu

Validation: Guo Tian, Haiyan Fu

Visualization: Xuelian Shi

Writing – original draft: Xuelian Shi

Writing – review & editing: Chunfu Wan, Xiaoli Xu

